# CTA versus TOF-MRA for circle of Willis segmentation: Implications for hemodynamic modelling

**DOI:** 10.64898/2026.04.10.26350583

**Authors:** A. Vikström, L. Zarrinkoob, M. Johannesdottir, A. Wåhlin, J. Hellström, M. Appelblad, P Holmlund

**Author notes:** Corresponding author: Axel Vikström, Department of Diagnostics and Intervention, Biomedical Engineering and Radiation Physics, Umeå University, S-901 87 Umeå, Sweden.

## Abstract

Modelling of hemodynamics in the circle of Willis (CoW) depends on vascular segmentation, which may vary based on imaging modality. Computed tomography angiography (CTA) is commonly used in clinic but involves radiation and injection of contrast agents, whereas magnetic resonance angiography (MRA) offers a non-invasive alternative. This study aims to compare CoW morphology and modelled cerebral perfusion pressure of CTA and MRA segmentations, validating if MRA can replace CTA in modelling workflows.

CTA and time-of-flight MRA (TOF-MRA) of the CoW was performed in 19 patients undergoing elective aortic arch surgery (67±7 years, 8 women). The CoW was semi-automatically segmented based on signal intensity thresholding. A TOF-MRA threshold was optimized against the CTA segmentation, using the CTA as reference standard. Computational fluid dynamics (CFD) modelling with boundary conditions based on subject-specific flow rates from 4D flow MRI simulated cerebral perfusion pressure in the segmented geometries. A baseline simulation and a unilateral brain inflow simulation, i.e., occlusion of a carotid, were carried out.

Linear mixed models indicated there was no effect of choice of modality on either average arterial lumen area (CTA – TOF-MRA: -0.2±1.3 mm^2^; p=0.762) or baseline pressure drops (0.2±1.9 mmHg; p=0.257). In the unilateral inflow simulation, we found no difference in pressure laterality (−6.6±18.4 mmHg; p=0.185) or collateral flow rate (10±46 ml/min; p=0.421).

TOF-MRA geometries can with signal intensity thresholding be matched to produce similar morphology and modelled cerebral perfusion pressure to CTA geometries. The modelled pressure drops over the collateral arteries were sensitive to the segmentation regardless of modality.

## Introduction

Hemodynamic modelling is emerging as a powerful approach for analyzing cerebral hemodynamics, i.e., cerebral perfusion pressure and blood flow [1]. Models integrate vascular geometries and flow data to simulate pressure and flow dynamics, providing insights into parameters that are difficult to measure directly. Modelling is therefore suitable for studying collateral circulation in the circle of Willis (CoW) [2]. The CoW helps maintain cerebral perfusion through alternative pathways in case of proximal stenosis or occlusion [3, 4]. While clinical imaging methods cannot predict the effects of steno-occlusive disease [5], modelling of blood flow in the CoW can help describe the collateral capacity for maintaining cerebral perfusion during such occlusions. In this context, cerebral perfusion pressure may serve as an indicator of the effective driving force for cerebral blood flow and potential hypoperfusion [6]. In recent development, the combination of vascular geometries segmented from computed tomography angiography (CTA) images and blood flow rates from 4D flow magnetic resonance imaging (4D flow MRI) with hemodynamic modelling has provided a method capable of assessing baseline cerebral perfusion pressure and cerebrovascular resistance in patients with carotid stenoses [7, 8]. The method has also demonstrated its feasibility in predicting laterality in cerebral perfusion pressure during different types of vascular surgery [9, 10]. However, the method’s current dependence on multiple image modalities, CTA and MRI, limits its clinical feasibility. Replacing the CTA with magnetic resonance angiography (MRA) and enabling the method to rely solely on MRI would also eliminate patient exposure to radiation and contrast agents.

The need for accurate vascular geometries implies model performance may be heavily influenced by the choice of image modality [11]. CTA and MRA are two modalities which have been shown to accurately visualize cerebral arteries, the CoW configuration and vascular pathology [12–15]. With the continued development of MRA sequences such as time-of-flight MRA (TOF-MRA), image characteristics like spatial resolution are improving to be on par with CTA [16]. However, despite these advances, no studies have thoroughly compared how CTA- and MRA-based segmentations affect hemodynamic outcomes in computational models of cerebral blood flow. Such investigations are especially important in the collateral circulation as small differences may generate substantial changes in pressure and flow resistance.

In this study, CTA-based segmentation of the CoW was used as reference to evaluate whether TOF-MRA can serve as a replacement in modelling of cerebral hemodynamics. We combined the segmentations with subject-specific flow data from 4D flow MRI in CFD simulations to determine whether baseline and occlusion-affected cerebral perfusion pressure can be predicted with a fully MRI-based approach.

## Methods

Three-dimensional geometries of the CoW were segmented from CTA and TOF-MRA brain images. The segmentation was semi-automatic and based on signal intensity thresholding, with the process kept consistent between the two modalities. Subject-specific blood flow rates were obtained from 4D flow MRI data. CFD simulations, using the segmented CoW geometries and blood flow rates, estimated cerebral hemodynamics at baseline and during virtual conditions of an occlusion in a carotid artery (i.e. unilateral brain inflow). Main parameters for comparison of the CTA and TOF-MRA geometries were average arterial lumen areas, simulated baseline pressure drops up to and within the CoW as well as simulated pressure laterality during unilateral carotid occlusion (Figure 1).

**Fig. 1.**
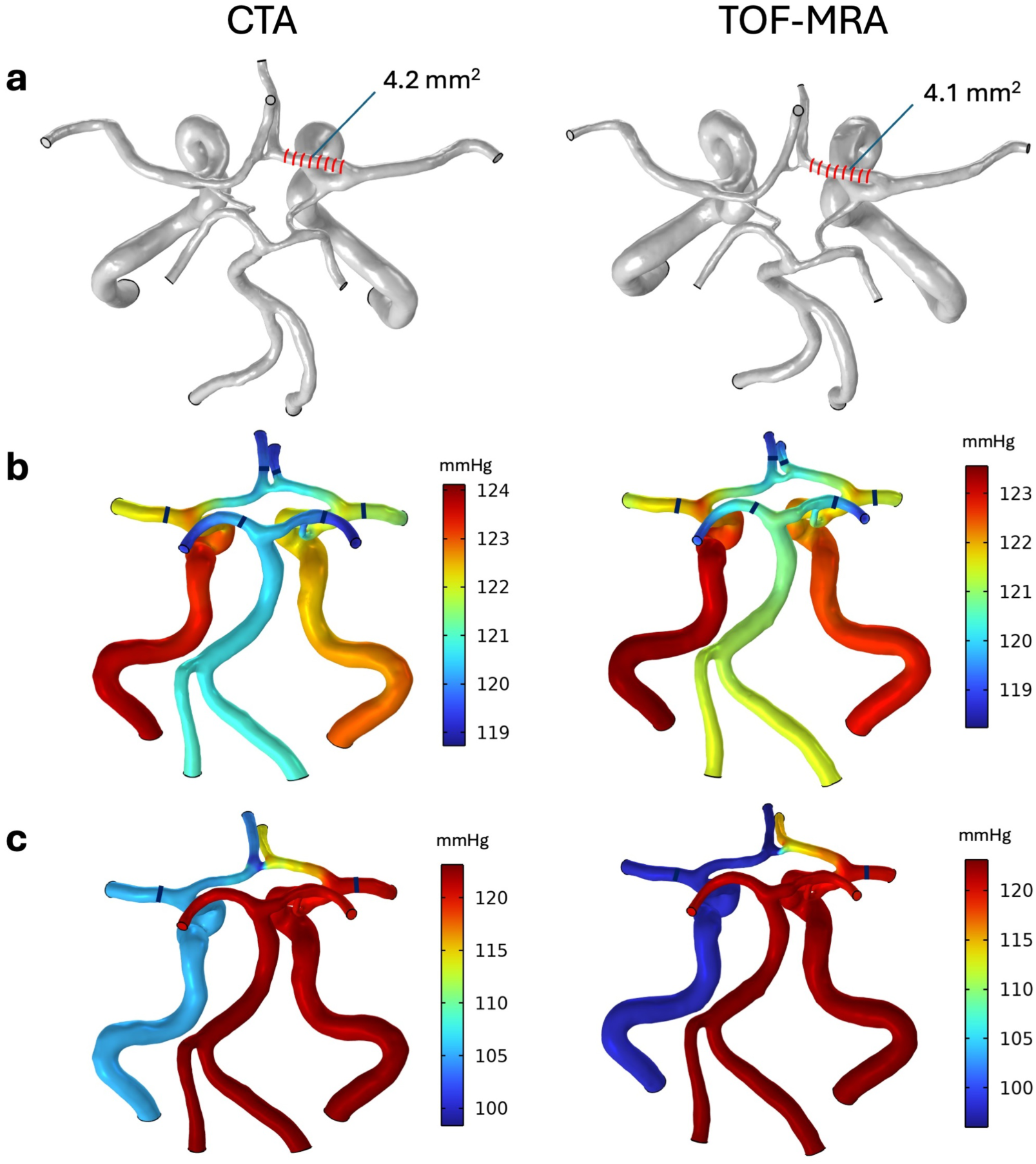
Segmented CoW geometries and modelled perfusion pressure distributions from the subsequent baseline and unilateral inflow simulations for both modalities. Note that the geometries in (a) belong to a different patient than the modelled pressure distributions. **(a)** Average arterial lumen areas were obtained from cross-sections along each artery. **(b)** Baseline pressure drops were computed as the difference in pressure between the inlets and the modelled perfusion pressure at each corresponding major cerebral artery. **(c)** Pressure laterality during unilateral inflow was computed as the difference in modelled perfusion pressure at the middle cerebral artery on the open and occluded side, here right and left side, respectively. The lines in the pressure distributions represents the positions at which the pressures were obtained from the model.

### Subjects

Patients undergoing elective aortic arch surgery at the Heart Centre, Cardiothoracic Surgery, Umeå University Hospital were recruited. Exclusion criteria were known contra-indication for MRI, previous stroke or kidney failure. Nineteen subjects were included; 8 women, age 67±7 years, systolic blood pressure (SBP) 160±27 mmHg, diastolic blood pressure (DBP) 96±16 mmHg, mean arterial pressure (MAP) 118±20 mmHg, heart rate 70±10 beats per minute.

The study was approved by the Swedish Ethical Review Authority (Dnr: 2022-02770-01) and performed in accordance with the guidelines of the Declaration of Helsinki. All participants were given oral and written information about the study and written consent was obtained from all participants.

### Imaging

CTA, MRA and 4D flow MRI data from head scans were acquired prior to surgery. CTA was conducted at each subject’s local hospital as part of the clinical routine. In-plane resolution was 0.5×0.5 mm^2^ and axial resolution varied between sites (0.3–0.625 mm^2^). MRI was performed with a 3T scanner (SIGNA™ Premier 3T MRI, GE Healthcare, Milwaukee, WI, USA) with a 48-channel head coil. MRA was performed with a 3D TOF-MRA sequence (Fat Sat, Hypersense, GE Healthcare), setup as: repetition time, 24 ms; echo time, 2.5 ms; flip angle, 15°; slice thickness, 1 mm; slice spacing, 0.5 mm; acquisition resolution, 400×400×64; imaging volume, 200×200×96 mm^3^; reconstructed resolution, 512×512×192; reconstructed voxel size, 0.39×0.39×0.50 mm^3^. Flow rates were acquired from a 4D flow MRI sequence utilizing a balanced 5-point phase contrast vastly undersampled isotropic projection reconstruction (PC-VIPR) sequence [17, 18]. Scan parameters were setup as: repetition time, 7.2 ms; echo time, 4.95 ms; velocity encoding, 110 cm/s; flip angle, 8°; radial projections, 16,000; acquisition resolution, 320×320×320; imaging volume, 220×220×220 mm^3^; reconstructed resolution, 320×320×320; reconstructed voxel size 0.7×0.7×0.7 mm^3^; 20 reconstructed cardiac time-frames. A non-invasive MRI-compatible blood pressure measuring device (CareTaker4, Caretaker Medical, Charlottesville, VA, USA) tracked the subject’s blood pressure with an inflatable finger cuff.

### Segmentation

The arterial geometries were semi-automatically segmented from the CTA and TOF-MRA datasets (Figure 2) using Synopsys’ Simpleware™ software (ScanIP P-2024.12; Synopsys, Inc., Mountain View, CA, USA). The segmentation included arteries of the CoW: internal carotid arteries (ICA), vertebral arteries (VA), the basilar artery (BA), the middle cerebral arteries (MCA1), proximal and distal posterior cerebral arteries (PCA1/PCA2), proximal and distal anterior cerebral arteries (ACA1/ACA2), and the anterior and posterior communicating arteries (ACoA and PCoA).

**Fig. 2.**
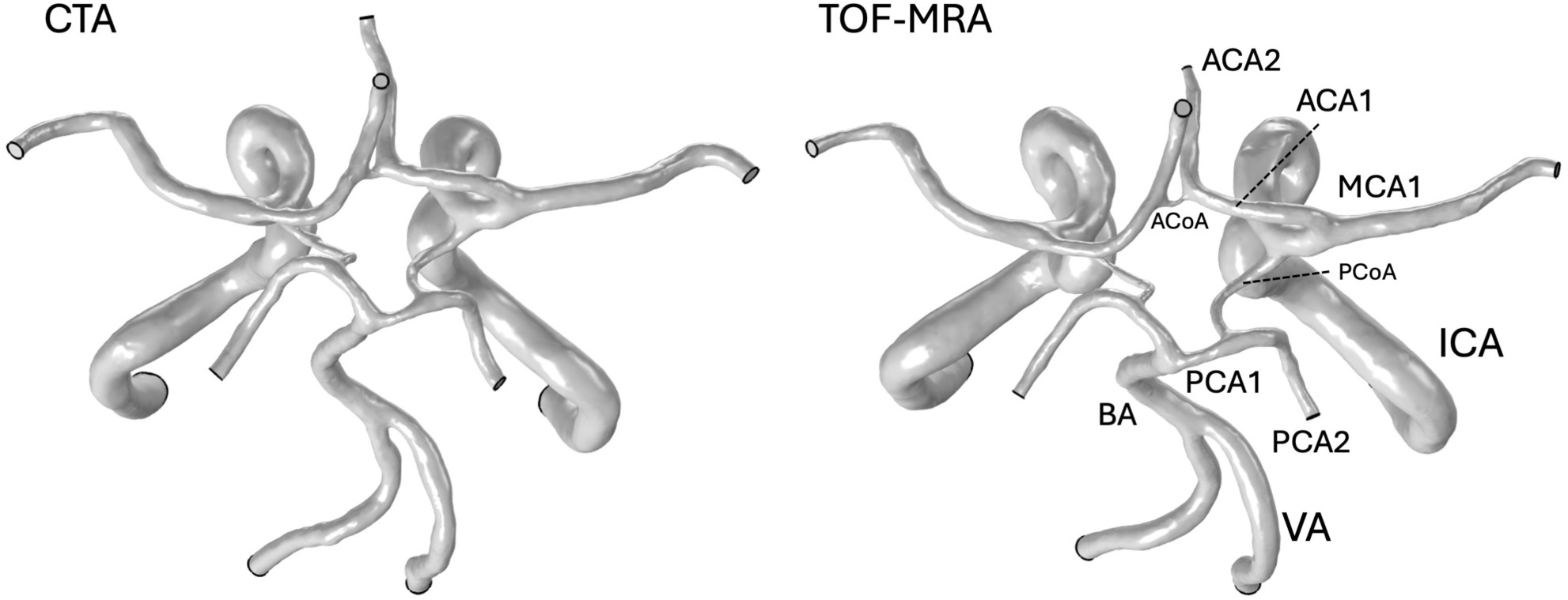
Segmentations of the CoW based on CTA and TOF-MRA data, cut off at the C2 segment.

Image pre-processing consisted of interpolating both the CTA and TOF-MRA data to isotropic voxels of 0.3×0.3×0.3 mm^3^, and bilateral filtering for reducing background noise. The original mask was established with signal intensity thresholding covering the signal range of the arteries. To simplify the segmentation, this was followed by a gradient-based filter from the software (*Local Surface Correction*, search radius: 2 voxels, smoothing radius: 2 voxels), which enhances separate tissues in the 3D workspace. Smaller arteries not of interest in this study were removed from the geometry. In the CTA segmentation, the ICAs and VAs had to be manually separated from interfering tissues like bone. The extracted vascular tree was then smoothed and dilated by one voxel in all directions, ensuring that the segmented mask encompassed approximately all signal for the arteries of interest. This step allowed a final round of thresholding restricted to the dilated mask and therefore only the vascular domain, preventing reintroduction of interfering tissues or unwanted minor arteries, while allowing the target arteries to be thresholded as intended (Figure 3).

**Fig. 3.**
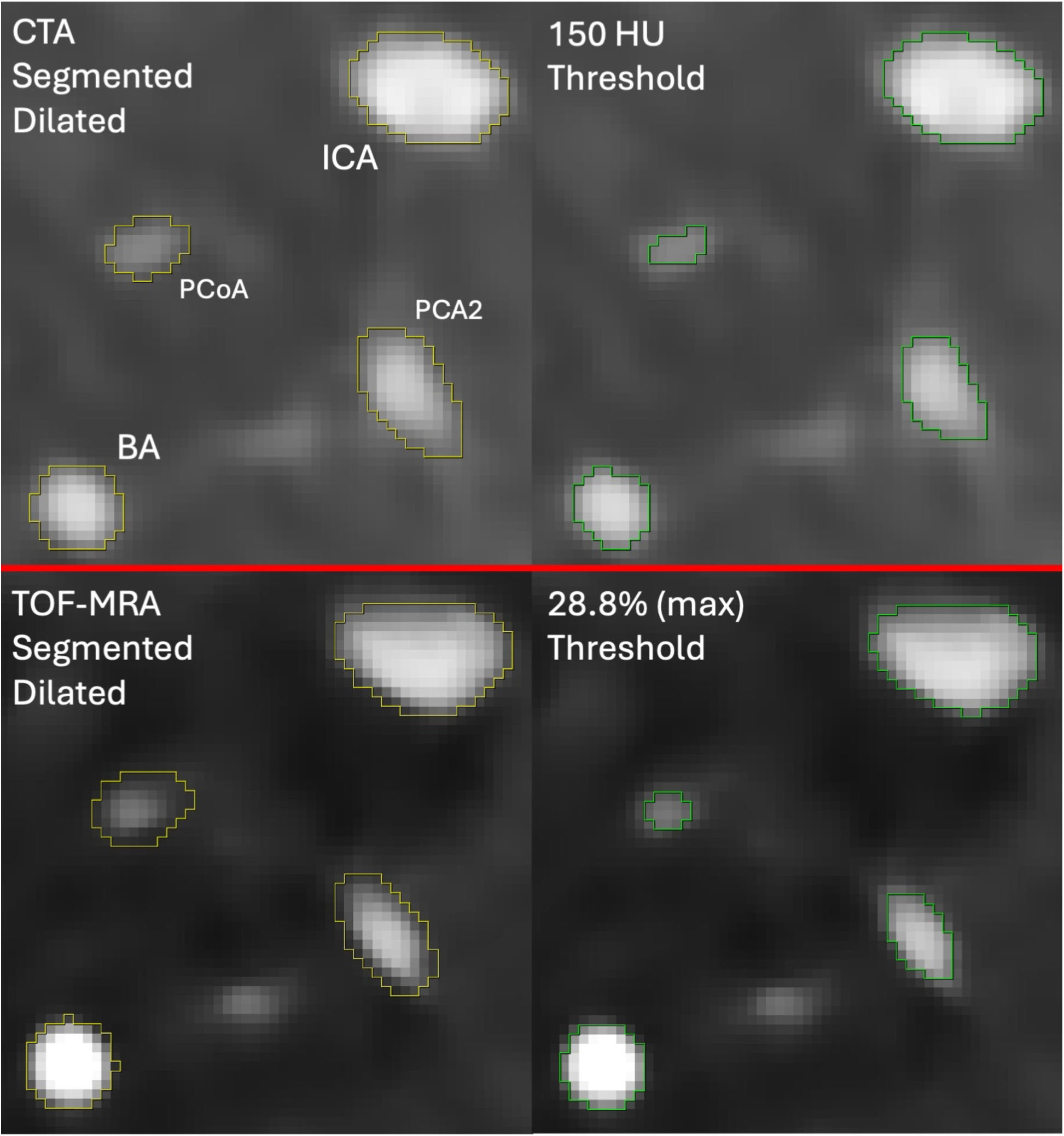
An axial slice in the CoW for both modalities. The yellow lines are the mask after smoothing and dilating of the vascular tree with excess arteries and interfering tissues removed. The green lines are the mask where voxels within the dilated mask (the vascular domain) have been thresholded with a lower limit of 150 HU in the CTA and 28.8% of the maximum intensity in TOF-MRA.

Previous works have identified 150 Hounsfield units (HU) as an appropriate lower threshold for CTA [19, 20]. The upper threshold limits the inclusion of tissue with higher signal than blood (e.g. bone) and was dependent on the largest arterial signal identified in the individual subjects, ranging between 450-600 HU. Only vascular signal is present in the TOF-MRA, implying that the lower threshold could be determined as a fraction of the maximum signal intensity while the upper threshold would correspond to the maximum signal intensity. Lower limits of 25% and 33.3% of the maximum intensity was expected to overestimate and underestimate the arterial lumen area, respectively, compared to the CTA (Online Resource 1). From these segmentations, we estimated through interpolation that 28.8% of the maximum intensity should yield the best overall arterial lumen area agreement with the CTA segmentations. Visible arteries with intensities slightly below the lower thresholds were for both modalities manually segmented with a lower threshold. This final mask was then smoothed with a volume and topology preserving smoothing filter in the software *(Recursive Gaussian*, 1 voxel). The vascular tree with the optimized threshold (28.8%) was used for all hemodynamic analysis of the TOF-MRA segmentations.

### Computational fluid dynamics

CFD models simulated pressure drops in the CoW, carried out in COMSOL Multiphysics® (COMSOL Multiphysics®, version 6.2, https://www.comsol.com, COMSOL AB, Stockholm, Sweden) by solving steady-state solutions to the Navier-Stokes equations. Laminar flow, blood as an incompressible Newtonian fluid and rigid arterial walls with a no-slip condition was assumed. Blood density was ρ = 1060 kg/m^3^ and viscosity was μ = 3.45 cP. Meshes were made with COMSOL’s in-built algorithm for fluid dynamics modelling, with 600,000 elements on average.

### Hemodynamic modelling of baseline and unilateral brain inflow

Two scenarios were simulated: one with measured baseline flows and one virtual setup with unilateral brain inflow corresponding to occlusion in a carotid (Figure 4).

**Fig. 4.**
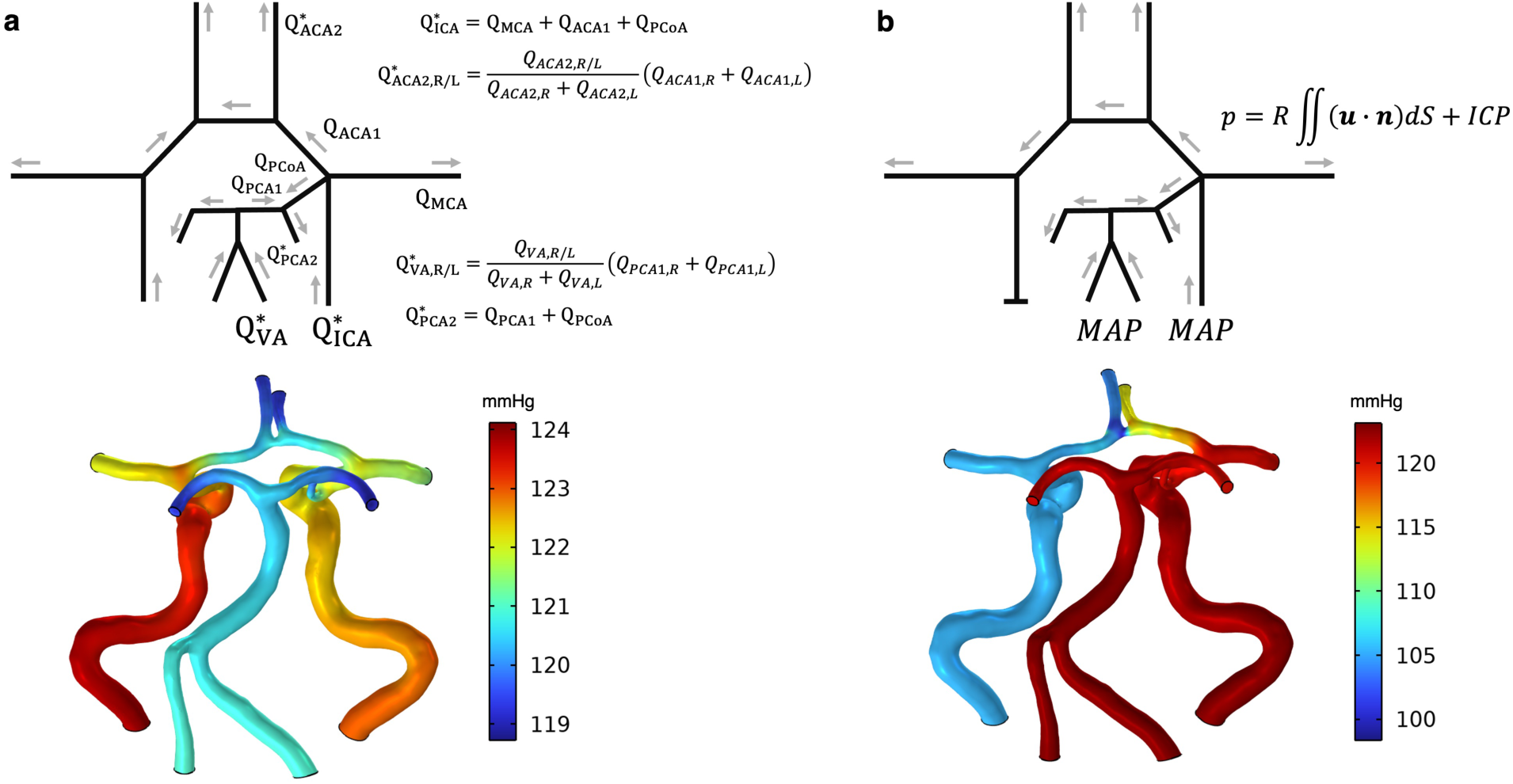
Simulation setup along with the simulated pressure distribution of the anterior circulation for a subject for **(a)** baseline and **(b)** virtual unilateral brain inflow. Flow rates 𝑄 were measured from subject-specific 4D flow MRI. The ACA2 flow rates were adjusted based on the ACA1 flows to preserve the measured flows in the CoW itself and while still achieving mass conservation. In the unilateral simulation, MAP was applied at the inflow boundaries whereas outflows were updated by the solver according to the expression for 𝑝. 𝑅 corresponds to the vascular resistance downstream of a cerebral artery to the parenchyma, determined from the computed perfusion pressures from the baseline simulation and the measured flow rates.

The baseline simulation was constructed using the flow rates in the CoW measured from the MRI (Figure 4a), with flow averaged over the cardiac cycle [21]. The ICAs and VAs were used as inlets, and the MCA1s, ACA2s and PCA2s were defined as outlets. Inflow rates were matched to the measured outflow rates to achieve mass conservation. Flow from the ACA1s to ACA2s does not exactly add up due to measurement uncertainties, which has been adjusted for by matching the ACA2 flows in the simulation based on the ACA1 flows [7]. The reference pressure of MAP was set as a pressure point condition on the ICA with the largest flow rate. If the geometry was disconnected by lacking PCoAs or ACoA, the reference pressure was also applied at the VA with the largest flow rate and the opposite ICA, respectively.

The unilateral inflow scenario was virtually constructed by completely removing the inflow from one ICA by adding a no-slip wall condition and letting pressure conditions govern the remaining inflows and outflows (Figure 4b). MAP was set as inflow boundary conditions at the open ICA. MAP was also applied at the VAs if the anterior and posterior circulations were connected by a PCoA, otherwise the posterior circulation was excluded from the simulation. Outflow boundary conditions were based on a previously described iterative method, where pressure is determined as the downstream territorial resistance 𝑅 times the flow rate through the artery, i.e. the pressure drop to the cerebral microcirculation, in addition to the intracranial pressure (ICP) [9, 10]. 𝑅 was for each cerebral artery computed from the baseline simulation as (𝑃_out_ − 𝐼𝐶𝑃)⁄𝑄_out_, using the predicted outlet pressure 𝑃_*out*_ and measured outflow 𝑄_*out*_. ICP was assumed to be 11.6 mmHg [22].

### Main comparative parameters

Average arterial lumen areas were in Simpleware extracted from the segmented geometries based on consecutive cross-sections orthogonal to the centerlines with 1.5 mm spacing. Areas of all included arteries except the exiting arteries, i.e. MCA1, ACA2 and PCA2, were included.

From the baseline simulation, pressure drops from the ICA and VA inlet pressure to five millimeters post-bifurcation on the corresponding MCA1, ACA2 and PCA2 arteries were obtained, corresponding to the cerebral perfusion pressure for the downstream vascular territories.

From the unilateral inflow simulation, the difference in perfusion pressure between the MCA1 on the side of the open ICA and the MCA1 on the side of the occluded ICA, i.e. the pressure laterality, was obtained. Moreover, the sum of the MCA1 and ACA2 outflows on the occluded side was obtained as a measure for the total collateral flow rate.

### Statistical analysis

All main parameters are presented as mean with standard deviation (mean±SD). P < 0.05 was the threshold for statistical significance. Normality was tested with the Lilliefors goodness-of-fit test.

Linear mixed models were made to account for multiple and different arteries in each subject in the analysis of arterial lumen areas and baseline pressure drops. For the morphological analysis, the model included arterial lumen area as dependent variable, modality and artery (intermediate arteries: ICA R/L, VA R/L, BA, ACA1 R/L, PCA1 R/L, PCoA R/L, ACoA) as fixed factor, modality and artery as an interaction and subject ID as random effect. In the baseline analysis, the model included logarithmized pressure drop as dependent variable, modality and artery (outflow arteries: MCA1 R/L, ACA2 R/L, PCA2 R/L) as fixed factors, modality and artery as an interaction and subject ID as random effect. The pressure drops had to be transformed, here logarithmized, to satisfy normality in the model residuals. Wilcoxon signed-rank test was used for paired comparison between the modalities for predicted MCA perfusion pressure laterality and collateral flow rate from the unilateral simulation, and correlation was investigated with Spearman’s rank correlation coefficient.

Bland-Altmann analysis was carried out for the arterial lumen areas and baseline pressure drops. As standard Bland-Altmann requires independent samples, the limits of agreement were computed with variances from linear mixed models of the difference between the modalities for these quantities. These models included difference in arterial lumen area or baseline pressure drops as dependent variable, artery as fixed factor (as above) and subject ID as random effect. The bias as well as between- and within-subject variances predicted by the model were used to compute the limits of agreement.

Lastly, intraclass correlation coefficient (ICC) analysis was carried out, comparing the TOF-MRA segmentations between two different raters using two-way mixed effects, absolute agreement, single measurement. All available arterial lumen areas, baseline pressure drops and pressure laterality for 10 random subjects were used in the comparison, resulting in 99 areas, 60 pressure drops and 7 pressure lateralities. The analysis was carried out per-artery and per-pressure drop as subjects contributed with dependent samples. Segmentation with CTA data has been tested in previous work, showing excellent interrater reliability [8]. The statistical analysis was carried out in MATLAB (R2024b, MathWorks, Natick, MA, USA) and jamovi (2.6.45.0, The jamovi project, Sydney, Australia).

## Results

### Morphological comparison

Vascular trees could be segmented from all subjects (N=19). There was excellent agreement in artery detection between the modalities, with the only exception being an ACoA visible in the CTA but not in the TOF-MRA for one subject. Three subjects lacked an ACoA and 15 subjects had no PCoA in both modalities. Inter-rater agreement for the TOF-MRA segmentation showed that ICC for all measured variables were excellent: the arterial lumen areas were all >0.99, ICC for baseline pressure drops were between [0.91–0.99] and pressure laterality had an ICC of 0.92 (CI 95%: [0.67–0.99]).

Arterial lumen areas from the CTA and TOF-MRA segmentations are illustrated in Figure 5, with average values given in Table 1. The linear mixed model indicated no effect of image modality on arterial lumen area (CTA – TOF-MRA; -0.2±1.3 mm^2^; p=0.762). The model presented conditional and marginal R^2^ of 0.90 and 0.86. Furthermore, the model indicated an effect of artery (p<0.001) but not for the artery-by-modality interaction (p=0.999).

**Fig. 5.**
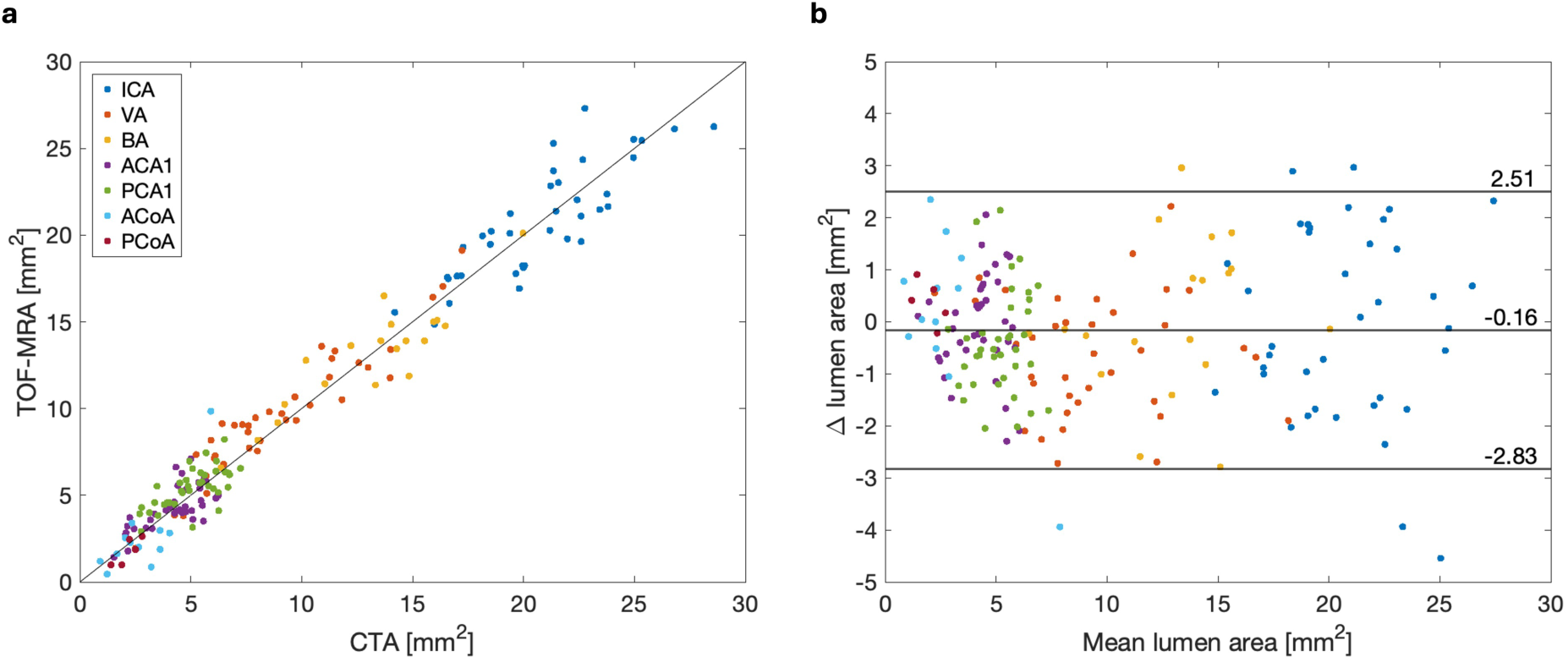
**(a)** Arterial lumen areas for the CTA and the optimized TOF-MRA geometries, for which choice of modality had no effect (p=0.762, linear mixed model). **(b)** Bland-Altmann plot with bias as well as limits of agreement determined from variances predicted by a linear mixed model of the difference in lumen area.

**Table 1.**
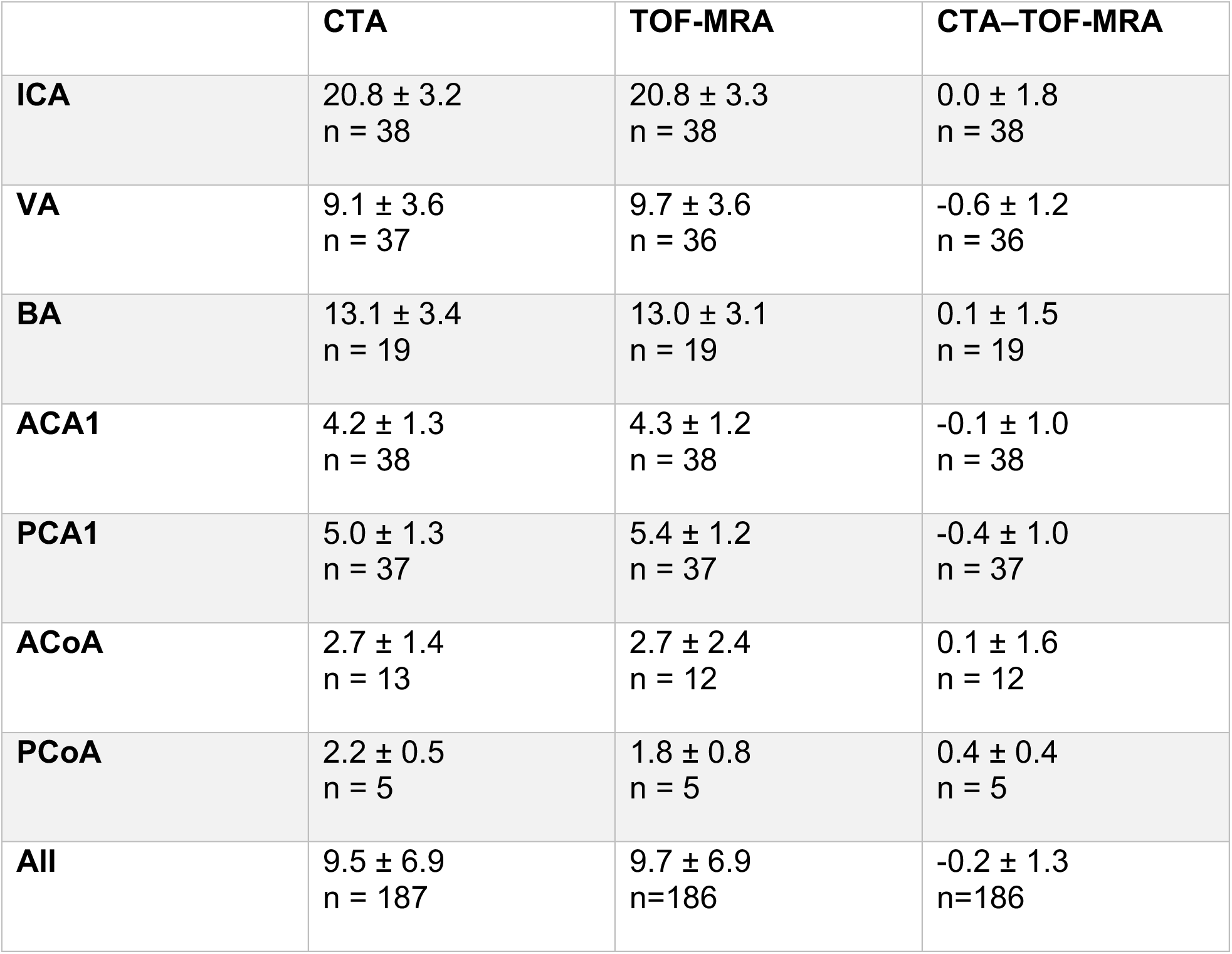
Average arterial lumen area (mean±SD) in mm^2^ for each modality and their difference.

### Hemodynamic comparison

Predicted pressure drops in the CoW during baseline are presented in Table 2 as well as Figure 6a and 6b. The linear mixed model indicated that the choice of modality did not have an effect on baseline pressure drops (CTA–TOF-MRA; 0.2±1.9 mmHg; p=0.257). The model presented conditional and marginal R^2^ of 0.82 and 0.68. There was an effect of artery (p<0.001) but not of the interaction between modality and artery (p=0.996).

**Table 2.** Pressure drops (mean±SD) in mmHg from the ICA/VA to the major cerebral artery outflows.

|  | **CTA** | **TOF-MRA** | **CTA-TOF-MRA** |
| --- | --- | --- | --- |
| **MCA1** | 2.4 ± 0.7<br>n = 38 | 2.4 ± 1.0<br>n = 38 | 0.0 ± 0.6<br>n = 38 |
| **ACA2** | 6.6 ± 3.8<br>n = 38 | 6.0 ± 3.4<br>n = 38 | 0.6 ± 3.1<br>n = 38 |
| **PCA2** | 1.5 ± 0.8<br>n = 38 | 1.4 ± 0.6<br>n = 38 | 0.1 ± 0.6<br>n = 38 |
| **All** | 3.5 ± 3.2<br>n = 114 | 3.3 ± 2.9<br>n = 114 | 0.2 ± 1.9<br>n = 114 |

**Fig. 6.**
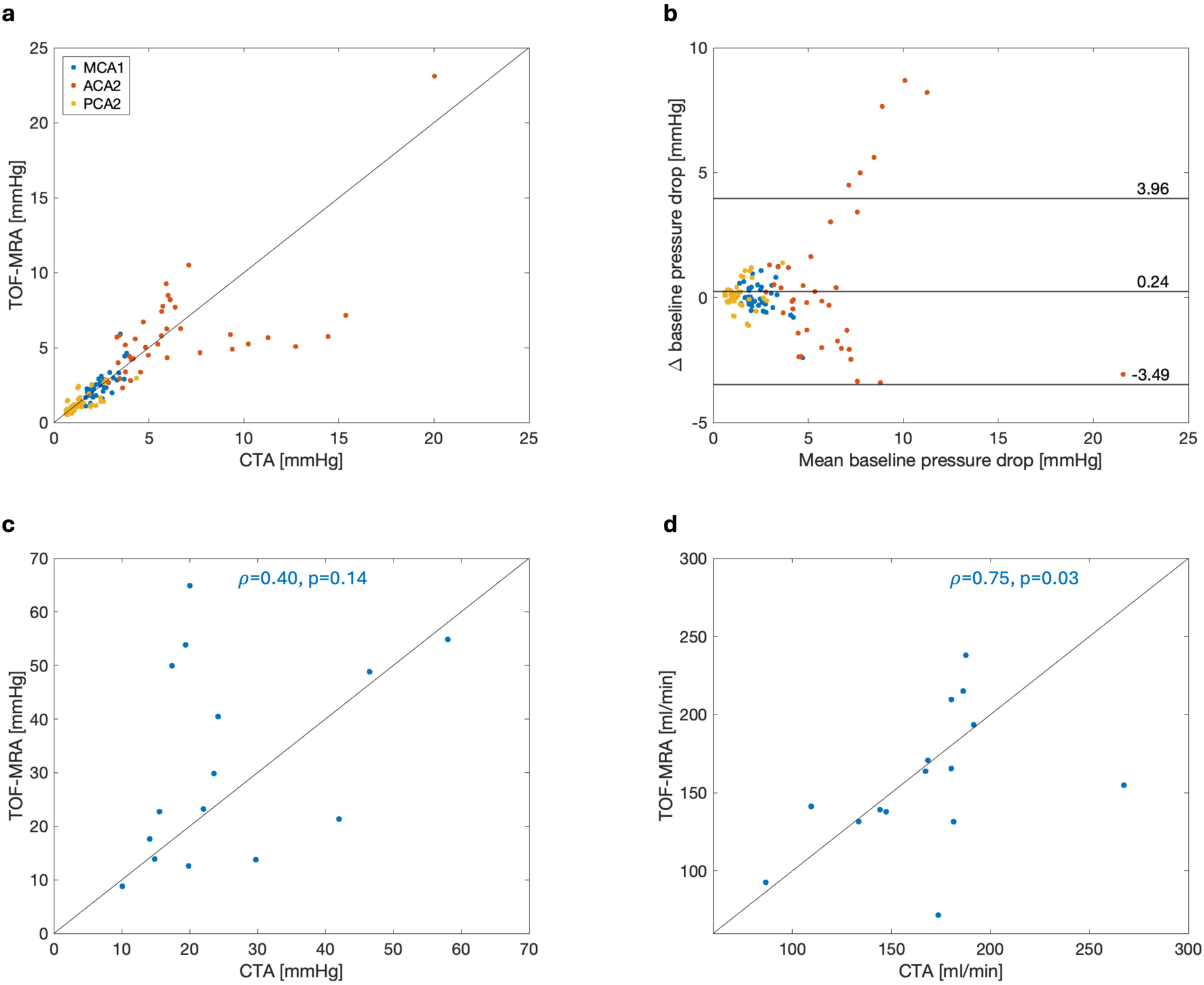
Results from the hemodynamic analysis of both modalities. (**a, b**) Predicted baseline pressure drops from ICA/VA to the cerebral arteries, for which choice of modality had no effect (p=0.257, linear mixed model). From the unilateral inflow simulation, **(c)** MCA1 pressure laterality did not differ between modalities (p=0.185, Wilcoxon signed-rank test) nor correlate (𝜌=0.40, p=0.14, Spearman). **(d)** Collateral flow rates did not differ (p=0.421, Wilcoxon signed-rank test) and were correlated (𝜌=0.57, p=0.03, Spearman).

The unilateral inflow simulation was carried out for 15 subjects as four subjects lacked an ACoA in either modality. This resulted in a predicted pressure laterality of 25.2 ± 13.5 mmHg for CTA and 31.8 ± 18.6 mmHg for TOF-MRA, without significant difference (−6.6±18.4 mmHg; p=0.185; Figure 6c). This difference in pressure laterality between the modalities ranged between -44.9–20.7 mmHg. Average collateral flow rate was 167.0 ± 41.3 ml/min for CTA and 157.1 ± 44.7 ml/min for TOF-MRA, also without significant difference (10±46 ml/min; p=0.421; Figure 6d). There was no correlation in predicted pressure laterality (𝜌 =0.40, p=0.14) whereas predicted collateral flow did correlate (𝜌=0.57, p=0.03).

## Discussion

In this study, we compared CoW morphology and modelled cerebral perfusion pressure of CTA and TOF-MRA geometries to validate if TOF-MRA can replace CTA in modelling workflows. With a segmentation method based on signal intensity thresholding and treating CTA as reference standard, an optimized TOF-MRA threshold was found such that morphology could be matched between the modalities. We did not find that choice of modality had an effect in the subsequent simulations of cerebral perfusion pressure. Our study shows that CTA and TOF-MRA can be used interchangeably for segmentation of the CoW, but that the modelling of pressure drops over the collateral arteries are sensitive to the segmentation for both modalities.

Morphology, in particular arterial size, dictates pressure drops in the vascular tree. Assessments of cerebral perfusion pressure are thus heavily dependent on the arterial lumen areas of segmented CoW geometries. Consequently, it is important that arterial lumen area is consistent between segmentations from both modalities. Position of the arterial wall is in this study determined by the lower threshold limit as the vascular signal attenuates outwards in the artery. By keeping the lower threshold for CTA constant at 150 HU [19, 20] and varying the lower TOF-MRA threshold based on a percentage of the maximum intensity in the image, we were able to identify 28.8% of TOF-MRA-maximum as the equivalent of 150 HU. The segmented arterial lumen areas therefore became highly similar between CTA and the optimized TOF-MRA geometries (Figure 5, Table 1). Compared to values from the literature, typically diameters [23, 24], the segmented areas reside above average but within presented ranges. It is also important to note that there was generally excellent agreement in amount of included arteries. One ACoA was present in CTA but not in the TOF-MRA for one subject which could be explained by TOF-MRA being flow dependent and that flow in this artery was insignificant at baseline.

There are several aspects of our semi-automatic signal intensity thresholding method that are critical for the resulting geometry. Firstly, the lower threshold is crucial and needs to be well defined. A high lower limit risks excluding arterial signal whereas a low lower limit may overestimate arterial signal by including partial volume effects. We therefore decided upon a literature-based lower CTA threshold of 150 HU [19, 20]. However, since the CTA data was obtained with scanners from different manufacturers, it could be that optimal threshold limits could vary among the patients. Secondly, signal may attenuate differently between modalities, and their representation of the artery may vary. With a real ground truth for arterial size in humans being unavailable besides imaging, the true position of the arterial wall within the vascular signal might differ between modalities. This is especially true in our case where one modality is contrast enhanced and the other is not, making it difficult to independently threshold and simultaneously match the modalities. We bypassed this by optimizing the TOF-MRA segmentation against the CTA. The interpolated TOF-MRA threshold worked remarkably well for all included arteries as the linear mixed model indicated no artery-by-modality interaction. This optimization approach should be feasible for TOF-MRA sequences with different acquisition parameters and possibly for other modalities. Lastly, this segmentation technique can be labor-intensive and operator-dependent as opposed to proposed automatic methods [11]. Automated segmentation methods are however often focused on creating a clinical tool for CoW variant classification rather than hemodynamic analysis and may underperform when anomalies like signal discontinuities are present [14]. The actual arterial size, except possibly identification of hypoplasia, is typically not investigated in such applications [25]. Thus, automated approaches could currently prove insufficient when accurate three-dimensional geometries are required [25]. Furthermore, ICC analysis demonstrated excellent operator independence for TOF-MRA segmentation, consistent with previous findings for CTA, indicating robust methodology [8].

Previous work comparing CTA and MRA in the context of hemodynamic analysis have studied vascular deformities such as aneurysms in both phantoms and humans, with varied conclusions [26, 27]. In humans, intracranial aneurysm morphology was similar between CTA and MRA while hemodynamic results were not [27]. Yet, in that study it was concluded that the general flow pattern in the surrounding vasculature should agree. We confirm this in our baseline simulations, as choice of modality had no effect on predicted pressure drops to each cerebral artery (Figure 6a, Table 2). This conclusion is mainly supported by the MCA1 and PCA2 pressure drops. However, there was a relatively large variation in the difference between modalities for the predicted pressure drops to the ACA2s (Figure 6b, Table 2). The difference in ACA1 arterial lumen area between the modalities presented a standard deviation of 1.0 mm^2^, corresponding to almost 25% of the average for both modalities. This variability combined with the relatively large flow rate for the average ACA1 size likely explains this spread in the pressure predictions. The spread in the pressure drops over the anterior collaterals was even more pronounced in the unilateral inflow scenario since it introduced substantial collateral flow over these arteries (Figure 6c). Yet, these results hints at the difficulty of segmenting the anterior collaterals in general rather than there being a difference between the modalities, given that there was no systematic difference in ACA1 arterial lumen area between the modalities.

Digital subtraction angiography (DSA) can be considered the gold standard for cerebrovascular imaging [28]. DSA is highly invasive and was not available for the included patients. Instead, CTA was in this study treated as the reference standard. This is partly due to its previous inclusion in the computational method, but also due to its well-established diagnostic use for vascular pathology. CTA does however also include a level of invasiveness due to the use of ionizing radiation and injection of contrast agents. Therefore, it would be beneficial to replace CTA with TOF-MRA in the modelling workflow. Our results support that CoW geometries segmented from TOF-MRA can be made comparable to CTA geometries in terms of morphology and modelled cerebral perfusion pressure. Moreover, the computational method’s current use of 4D flow MRI implies that the method would logistically benefit from an MRA-dependent segmentation, as the amount of imaging sessions and modalities would be reduced.

This study presents some limitations. A major limitation is that we have no ground truth for either the arterial lumen areas or the baseline pressure drops. However, the aim was to optimize and compare the TOF-MRA geometries against the CTA geometries, implying that the true values are not necessary for the actual comparison. Validation of the geometries must in turn be carried out in the subsequent modelling by reproducing measured hemodynamic parameters, e.g. intraoperative blood pressure measurements [9, 10]. Another limitation is that while the CTA threshold of 150 HU is more general, there is limited generalizability of the presented TOF-MRA threshold of 28.8% considering the signal intensity will likely vary depending on the TOF-MRA acquisition. Our optimization approach should however prove sufficient to find a threshold for other MRA sequences or modalities. There was also inherent variability in the CTA images as they were collected at each subject’s hospital, while the TOF-MRA was carried out at the same MRI scanner.

This study presents a morphological and hemodynamic comparison of arterial geometries of the CoW segmented from CTA and TOF-MRA images. Using signal intensity thresholding, an appropriate threshold for TOF-MRA could be found such that average arterial lumen area for the included arteries agreed well between the modalities. Subsequent simulations generated cerebral perfusion pressures that were similar, however the modelled pressure drop over the anterior collaterals was for both modalities likely sensitive to the segmentation. Overall, the results suggest that TOF-MRA is a suitable alternative to CTA for segmentation of the CoW.

## Declarations

### Ethics approval

The study was approved by the Swedish Ethical Review Authority (Dnr: 2022-02770-01) and performed in accordance with the guidelines of the Declaration of Helsinki.

### Consent

All participants were given oral and written information about the study and written consent was obtained from all participants.

### Data availability

Data is available from the corresponding authors upon reasonable request.

## Author contributions

**Axel Vikström:** Conceptualization, Methodology, Software, Validation, Formal Analysis, Investigation, Data Curation, Writing – Original Draft, Writing – Review & Editing, Visualization. **Laleh Zarrinkoob:** Investigation, Writing – Review & Editing. **Martha Johannesdottir:** Investigation, Resources, Writing – Review & Editing. **Anders Wåhlin:** Investigation, Writing – Review & Editing. **Jan Hellström:** Investigation, Resources, Writing – Review & Editing, Project Administration. **Micael Appelblad:** Investigation, Resources, Writing – Review & Editing. **Petter Holmlund:** Conceptualization, Methodology, Validation, Investigation, Resources, Data Curation, Writing – Review & Editing, Supervision, Project Administration.

## Statements and Declarations

The authors have no competing interests to declare that are relevant to the content of this article.

## Data availability statement

Data is available from the corresponding authors upon reasonable request.

## Supporting information

Online Resource 1

## Acknowledgements

The project was supported by Region Västerbotten through Centrala ALF funding and Spjutspetsmedel. Study sponsors had no involvement in the study.

