## Supplementary material for "CTA versus TOF-MRA for circle of Willis segmentation: Implications for hemodynamic modelling": Online Resource 1

**Journal**: Annals of Biomedical Engineering

**Authors**: Vikström, A.^1*^, Zarrinkoob, L.^2^, Johannesdottir, M.^3^, Wåhlin, A.^4,5^, Hellström, J.^3^, Appelblad, M.^3^, Holmlund, P^4^.

**Affiliations**: ^1^ Department of Diagnostics and Intervention, Biomedical Engineering and Radiation Physics, Umeå University, Umeå, Sweden

^2^ Department of Diagnostics and Intervention, Anesthesiology and Intensive Care, Umeå University, Umeå, Sweden

^3^ Department of Public Health and Clinical Medicine – Heart Centre, Umeå University, Umeå, Sweden

^4^ Department of Applied Physics and Electronics, Umeå University, Umeå, Sweden

^5^ Umeå Center for Functional Brain Imaging, Umeå University, Umeå, Sweden

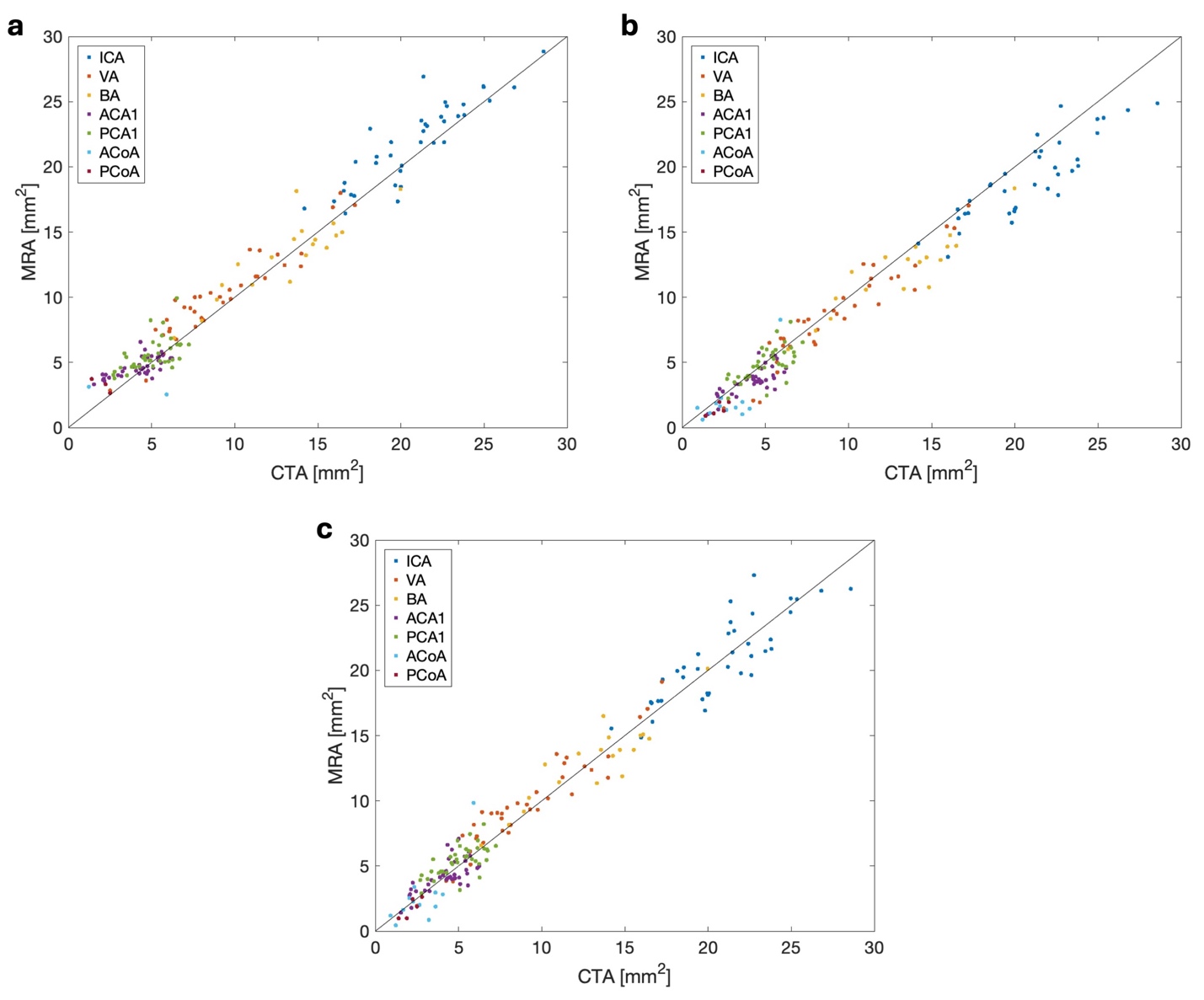

**Fig. 1** Arterial lumen areas for the arteries for the CTA segmentation (150 HU) and the TOF-MRA thresholds based on **(a)** 25.0%, **(b)** 33.3% and **(c)** 28.8% of the image maximum intensity.
